# Prospective Bidirectional Associations Between Core Schemas, Attenuated Psychotic Symptoms, and Functioning in Youth at Clinical High Risk for Psychosis

**DOI:** 10.64898/2026.09.30.26364206

**Authors:** Alexander Broekhuijse, Abhishek Saxena, Catalina Mourgues-Codern, Hesham Mukhtar, Jean Addington, Carrie E. Bearden, Kristin S. Cadenhead, Daniel H. Mathalon, Thomas H. McGlashan, Diana O. Perkins, Elaine F. Walker, Tyrone D. Cannon, Scott W. Woods, Albert R. Powers, Emily A. Farina

## Abstract

**Objective:** Cognitive models of psychosis implicate maladaptive core schemas in the development and emergence of psychotic symptoms, but no study has tested temporal dynamics between schemas and psychotic symptoms in a large sample of clinical high-risk (CHR) youth. Given their sizable role in cognitive behavioral therapies, the first-line treatment for CHR youth, core schemas may be key to both symptom development and treatment in this population.

**Methods:** Participants were 578 CHR individuals from the second North American Prodrome Longitudinal Study (NAPLS-2) assessed across five timepoints. Time-lagged mixed-effects models tested forward (schema→outcome) and reverse (outcome→schema) paths for each Brief Core Schema Scales subscale with both psychotic symptom and functioning changes, controlling for autoregressive effects, time-varying depression, and linear time.

**Results:** Twenty of 36 schema–outcome pairs showed significant prospective associations. Negative-other schemas were prospectively associated with worsening across nearly all psychosis symptom and functioning domains. Suspiciousness exhibited bidirectional associations with all four schema subscales. Grandiose ideas were uniquely prospectively associated with positive-self and negative-other schemas. Positive schemas were prospectively associated with decreases in suspiciousness, negative symptoms, and improvements in functioning. Effects were largely independent of depressive symptoms and robust across sensitivity analyses, with partial attenuation of self-schema effects on suspiciousness.

**Conclusions:** Core schemas were prospectively associated with psychosis symptom and functioning change in CHR youth, with symptom-domain specificity and bidirectionality. These findings extend cognitive models of psychosis into CHR syndromes and highlight schemas as potentially modifiable treatment targets to improve outcomes in this population.

**Highlights:**

- Schema changes preceded shifts in functioning and psychosis symptoms among CHR youth
- Negative-other schemas preceded worsening across nearly all outcome domains
- Suspiciousness was the only outcome with bidirectional schema associations
- Positive schemas preceded fewer negative symptoms and better global functioning

## Introduction

While outcomes for young people at clinical high risk for psychosis (CHR) are diverse, many individuals experience long-term functional decline, persistent attenuated positive symptoms, and numerous comorbidities, even amongst those who do not transition to psychosis (Fusar-Poli et al., 2015; Rutigliano et al., 2016). Existing psychosocial interventions for CHR show inconsistent effects on transition, do not influence functioning compared to control conditions, and have only small effects on attenuated positive symptoms (pooled SMD −0.15 to −0.24 at 12– 24 months; Fusar-Poli, 2019; Mei, 2021; Zheng et al., 2022), signaling the need to identify modifiable treatment targets prospectively associated with psychotic symptom emergence and functional decline. Core schemas are widely regarded as transdiagnostic mechanisms of change in cognitive behavioral therapies (Bär et al., 2023; Beck and Haigh, 2014; Sunde et al., 2019; Van Tatenhove et al., 2025; Wegener et al., 2013), the first-line treatment for CHR youth (Poletti et al., 2024), making them candidate treatment targets if their prospective role in CHR symptom dynamics can be established.

In the psychosis literature, core schemas are defined as global evaluative beliefs about oneself and others (Garety et al., 2001). These schemas are formed by early experience and subsequently influence the appraisal of interpersonal, affective, and sensory stimuli (Beck and Rector, 2005; Freeman et al., 2002; Garety et al., 2001). Fowler et al. (2006) operationalized this construct with the Brief Core Schema Scales (BCSS), and this term has been used across the psychosis continuum (Smith et al., 2006; Taylor et al., 2014) and in CHR samples (Addington and Tran, 2009; Stowkowy et al., 2016; Stowkowy and Addington, 2012). This deviates from the definition of schemas used in schema therapy, which defines schemas as pervasive patterns of memories, emotions, cognitions, and bodily sensations organized around specific themes including mistrust/abuse (Young, 2006). Instead, this definition more closely aligns with the concept of core schemas introduced in Beck and Rector’s (2005) cognitive therapy of schizophrenia, and core beliefs in traditional Cognitive Therapy models (Beck and Haigh, 2014; Jorovat et al., 2025). We retain the term core schemas to maintain consistency with the instrument used and the foundational CHR literature.

Cognitive models of psychosis development (Beck and Rector, 2005; Freeman et al., 2002; Garety et al., 2007, 2001) suggest that core schemas bias one’s interpretation of anomalous experiences leading to the development and reinforcement of psychotic symptoms. Two features of these models have received limited empirical attention in CHR populations. First, the models imply symptom-domain specificity: negative self- and other-schemas drive threat-based appraisals that generate persecutory ideation, unusual thought content, and distressing interpretations of perceptual abnormalities. Alternatively, other pathways may underlie symptoms such as grandiose ideas, where inflated self-appraisal rather than threat may drive symptom severity. Second, the models posit bidirectionality, wherein schemas shape symptom appraisal but symptoms, once established, reinforce the original schemas (Freeman et al., 2002).

Longitudinal models of psychosis development further implicate core schemas. The biopsychosocial model of transition (Georgiades et al., 2023) proposes that current psychosocial stress activates pre-existing negative schemas, worsening affect and reinforcing reasoning biases, prompting a misinterpretation of anomalous stimuli. The affective pathway model (Gurnani and Georgiades, 2025) positions core schemas as a conduit between interpersonal adversity and negative affect, ultimately leading to psychotic symptoms (Jaya et al., 2018). Similarly, core schemas have been proposed to mediate the effects of trauma and bullying on attenuated psychotic symptoms (Jorovat et al., 2025; Stowkowy and Addington, 2012). However, these models were developed in individuals with established psychotic disorders or from cross-sectional CHR data, and their temporal claims have not been confirmed longitudinally in CHR samples.

Emerging research has attempted to explore these processes in CHR youth, with cross-sectional studies finding that CHR individuals endorse more negative and fewer positive schemas than controls (Addington and Tran, 2009; Jorovat et al., 2025; Stowkowy et al., 2016). Preliminary longitudinal work suggests that schema changes may predict changes in symptom severity and functioning, though these findings were underpowered and inconclusive (Patton et al., 2022).

Notably, no study has tested the temporal dynamics suggested by cognitive models within CHR youth. Additionally, past research has examined positive symptoms as a unitary construct, obscuring the possibility that different symptoms have distinct schema associations. Positive schemas have been comparatively neglected (Bemrose et al., 2021), despite meta-analytic evidence that CHR populations have fewer positive beliefs (Jorovat et al., 2025) and the possibility that they may be protective. Existing evidence therefore cannot evaluate the temporal, specific, or bidirectional claims cognitive models posit, nor speak to schemas as resilience factors, limiting their translation into intervention targets for CHR youth.

The present study utilized time-lagged mixed effects models in a large multisite CHR cohort (North American Prodrome Longitudinal Study – 2, NAPLS-2, N=764, Addington et al., 2015) to examine prospective associations between positive and negative self and other schemas and individual positive symptom domains, total negative symptoms, and functioning across five timepoints. Analyses tested both forward paths (schema predicting subsequent symptom/functioning change) and reverse paths (symptom/functioning predicting subsequent schema change) to evaluate bidirectionality. We hypothesized that (1) negative-self and negative-other schemas would be prospectively associated with increases in unusual thought content, suspiciousness, and perceptual abnormalities, consistent with the threat-appraisal pathway (Garety et al., 2001); (2) suspiciousness would show distinct bidirectional associations with schemas, reflecting the mutual reinforcement posited by threat-anticipation models (Freeman et al., 2002); (3) grandiose ideas would be associated with positive-self rather than negative schemas, consistent with inflated rather than threat-based appraisal; and (4) positive schemas would be prospectively associated with better functioning and lower psychosis symptom burden. This design provides the first large-scale test of whether schema dynamics operate prospectively, bidirectionally, and with symptom-domain specificity in youth at CHR.

## Methods

### Participants

The sample comprised CHR participants recruited through the second wave of the North American Prodrome Longitudinal Study (NAPLS-2) (Addington et al., 2015). All participants completed written informed consent after receiving complete descriptions of the constituent studies, which were approved by local ethics committees. Recruitment and procedures are described in detail elsewhere (Addington et al., 2015). The NAPLS-2 cohort was composed of 764 individuals who met the criteria of prodromal symptoms detailed in the SIPS. Those with at least two BCSS timepoints were included in analyses.

### Measures

Demographic, functional, symptomatic, and cognitive-schema variables assessed in NAPLS-2 were included in analyses. All assessments have documented validity for youth at CHR and were utilized by trained raters who confirmed diagnostic decisions at weekly consensus calls.

The Brief Core Schema Scales (BCSS) is a 24-item self-report measure assessing beliefs about the self and others on a five-point rating scale, with higher scores indicating greater agreement with general statements about oneself (“I am worthless”) and others (“Other people are unforgiving”; scored 0-4; Taylor et al., 2014). Four schema scores are produced: negative beliefs about oneself (negative-self), negative beliefs about others (negative-others), positive beliefs about the self (positive-self), and positive beliefs about others (positive-others) (range 0 to 24). This scale has demonstrated validity in the CHR population (Addington and Tran, 2009).

The Structured Interview for Psychosis-risk Syndromes (SIPS; Miller et al., 2003) is a structured clinical interview composed of four scales that measures positive symptoms (range 0-30), negative symptoms (range 0-36), disorganization (range 0-24), and general psychopathology (range 0-24). Each item is scored on a scale of 0 (Absent) to 6 (Psychotic), with a 6 indicating the presence of frank psychosis and 3-5 indicating the CHR severity range. Positive symptom items include unusual thought content/delusional ideas (P1), suspiciousness/persecutory ideas (P2), grandiose ideas (P3), perceptual abnormalities/hallucinations (P4), and disorganized communication (P5). Symptom scores were verified through weekly consensus calls and evaluations were performed by certified raters.

The Calgary Depression Scale for Schizophrenia (CDSS) is a nine item, clinician-administered interview developed to assess depressive symptoms in individuals with psychotic disorders, including those at CHR (Addington et al., 1993, 2014). Items (depression, hopelessness, self-deprecation, guilty ideas of reference, pathological guilt, morning depression, early awakening, suicide, observed depression) are rated on a scale of zero to three and summed to develop a total score (range 0–27; higher scores = greater depression).

The Global Assessment of Functioning (GAF) is a single-item clinician rating of overall psychological, social, and occupational functioning on a 0 to 100 scale, anchored in ten-point intervals (Endicott, 1976).

The Global Functioning: Social scale (GFS; Cornblatt et al., 2007) is a single-item clinician-rated measure of social functioning on a ten-point scale which assesses quantity and quality of peer relationships, interpersonal conflict, and age-appropriate social engagement. The Global Functioning: Role scale (Cornblatt et al., 2007) measures performance in work, school, or homemaker roles relative to age-appropriate standards.

### Statistical Analyses

Statistical analyses were conducted in R version 4.5.1 with the *lme4* package (Bates et al., 2015). To examine prospective associations between changes in cognitive schemas and changes in positive and negative symptom severity and functioning over time, we estimated time-lagged mixed-effects models predicting each outcome at time *t* from each schema at time *t-1*, controlling for the autoregressive effect of the outcome at *t-1*, time-varying depression, and linear time.

Time-varying depression (CDSS total score at each assessment) was included as a covariate in all models to account for the well-documented co-occurrence of depressive symptoms and negative schematic beliefs in CHR populations (Smith et al., 2006) and to provide a conservative test of whether schema effects on clinical outcomes are independent of concurrent mood state.

Parallel models tested reverse paths (outcome at *t-1* predicting schema at *t*). All models included participant ID as a random intercept. All variables were standardized prior to model estimation. Analyses used all available observations; participants contributed data to a given model if they had non-missing values on the relevant schema and outcome at consecutive timepoints. False Discovery Rate (FDR) correction was applied across all 36 schema-outcome pairs, with corrected *p*<0.05 considered significant (q). Directionality was inferred when the forward path was significant and the reverse path was not, bidirectional associations when both paths were significant.

We estimated prospective schema–outcome associations using time-lagged mixed-effects models with a participant random intercept and an autoregressive term. While a random-intercept cross-lagged panel model (RI-CLPM; Hamaker et al., 2015) separates within-from between-person variance more cleanly, identification of a stable-trait component requires at least three assessments of each construct, and only 275 participants (36.0%) completed four or more BCSS administrations. Estimating multi-pathway latent specifications stably across 72 models with time varying covariates was therefore not feasible. Time-lagged mixed-effects models accommodate unbalanced and partially missing data through maximum likelihood and fit uniformly across all pairs. To address potential conflation of within- and between-person variance, we report intraclass correlations for every variable and refit all significant models with a random slope for time to test whether individual differences in trajectory altered the estimates; coefficients changed by |Δβ| ≤ .007 (**Table S5**). Following recommendation against applying any single cross-lagged framework by default (Lüdtke and Robitzsch, 2022; Orth et al., 2021), we selected the model best aligned with our data structure and research questions.

## Results

### Sample and model characteristics

The analytic sample included 578 participants with at least two BCSS assessments across five timepoints at eight sites. These participants had mean schema scores of 6.56 (Negative-self), 10.95 (Positive-self), 7.92 (Negative-other), and 9.64 (Positive-other). Due to missing data on schema measures, individual time-lagged models included 1,279–1,291 observations from 518– 521 unique participants. Additional details on the analytic sample are available in **Table 1**.

**Table 1.**
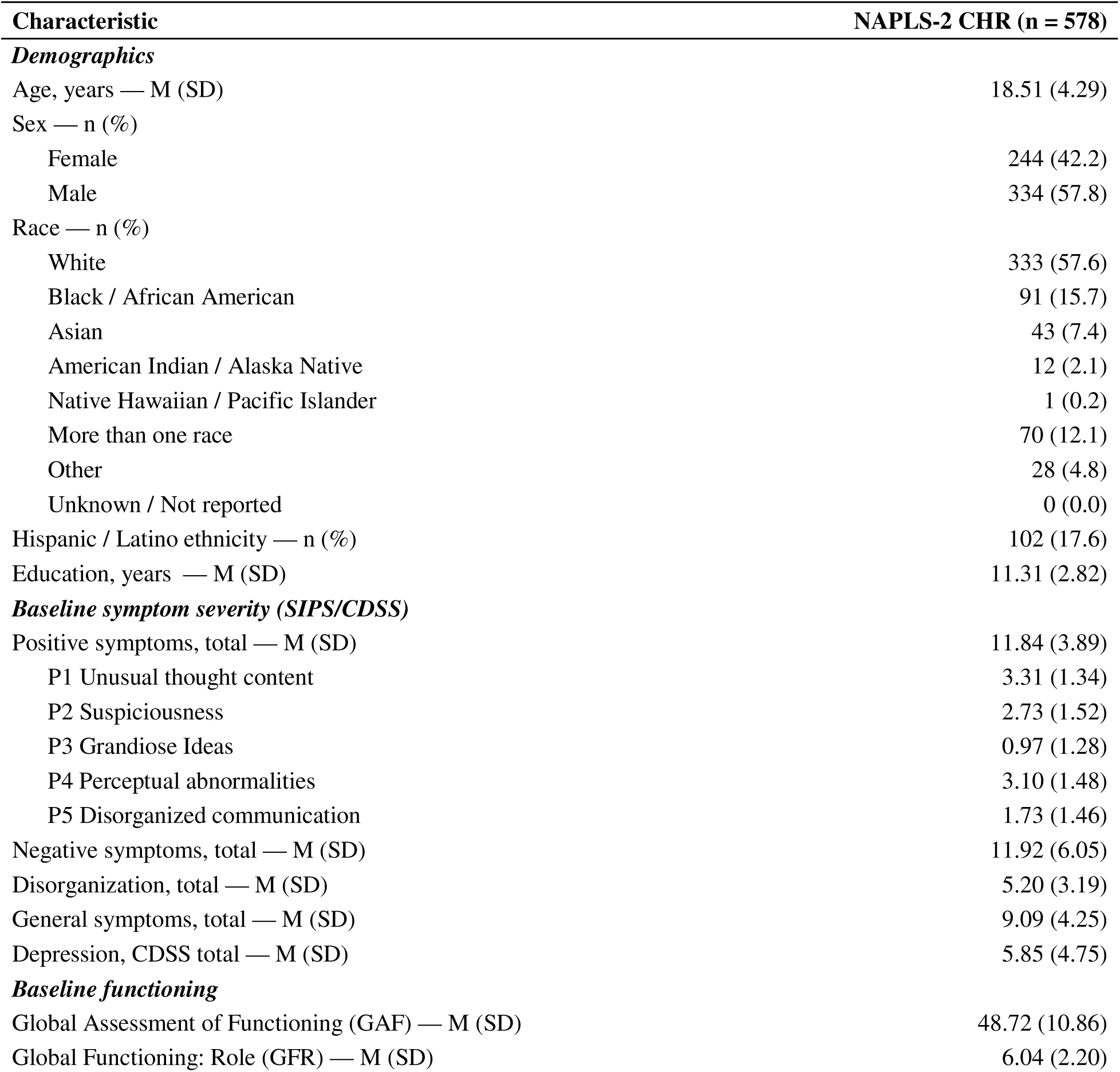
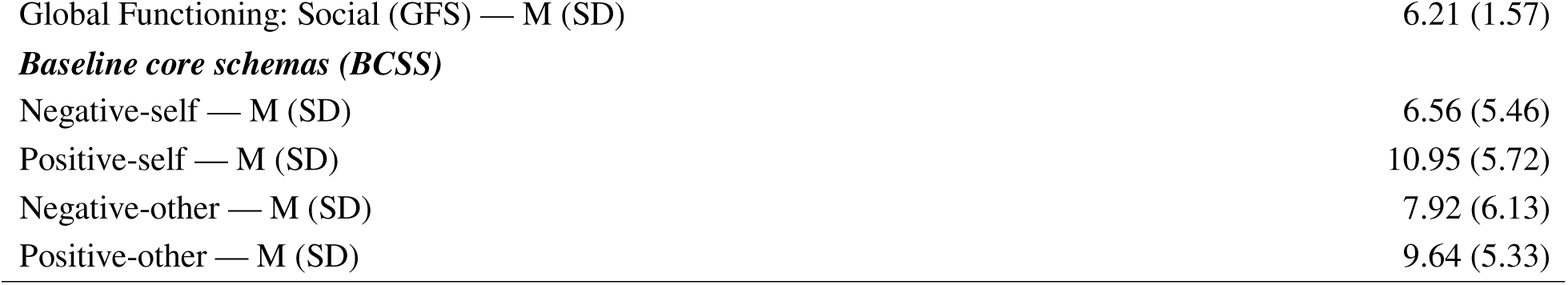
Baseline demographic and clinical characteristics of the NAPLS-2 CHR participants with ≥ 2 BCSS assessments.

| **Characteristic** | **NAPLS-2 CHR (n = 578)** |
| --- | --- |
| ***Demographics*** |  |
| Age, years — M (SD) | 18.51 (4.29) |
| Sex — n (%) |  |
| Female | 244 (42.2) |
| Male | 334 (57.8) |
| Race — n (%) |  |
| White | 333 (57.6) |
| Black / African American | 91 (15.7) |
| Asian | 43 (7.4) |
| American Indian / Alaska Native | 12 (2.1) |
| Native Hawaiian / Pacific Islander | 1 (0.2) |
| More than one race | 70 (12.1) |
| Other | 28 (4.8) |
| Unknown / Not reported | 0 (0.0) |
| Hispanic / Latino ethnicity — n (%) | 102 (17.6) |
| Education, years — M (SD) | 11.31 (2.82) |
| ***Baseline symptom severity (SIPS/CDSS)*** |  |
| Positive symptoms, total — M (SD) | 11.84 (3.89) |
| P1 Unusual thought content | 3.31 (1.34) |
| P2 Suspiciousness | 2.73 (1.52) |
| P3 Grandiose Ideas | 0.97 (1.28) |
| P4 Perceptual abnormalities | 3.10 (1.48) |
| P5 Disorganized communication | 1.73 (1.46) |
| Negative symptoms, total — M (SD) | 11.92 (6.05) |
| Disorganization, total — M (SD) | 5.20 (3.19) |
| General symptoms, total — M (SD) | 9.09 (4.25) |
| Depression, CDSS total — M (SD) | 5.85 (4.75) |
| ***Baseline functioning*** |  |
| Global Assessment of Functioning (GAF) — M (SD) | 48.72 (10.86) |
| Global Functioning: Role (GFR) — M (SD) | 6.04 (2.20) |
| Global Functioning: Social (GFS) — M (SD) | 6.21 (1.57) |
| **Baseline core schemas (BCSS)** |  |
| Negative-self — M (SD) | 6.56 (5.46) |
| Positive-self — M (SD) | 10.95 (5.72) |
| Negative-other — M (SD) | 7.92 (6.13) |
| Positive-other — M (SD) | 9.64 (5.33) |

Intraclass correlation coefficients (ICC) indicated between-person differences accounted for approximately half of the variance across all variable types (schemas mean ICC=.56; symptoms mean ICC=.53; functioning mean ICC=.55), confirming substantial within-person variability suitable for time-lagged mixed effect modeling. Autoregressive coefficients were moderate to high for all outcomes (symptoms: mean β = .57, range = .47–.69; functioning: mean β = .59, range = .45–.71; variable specific ICCs available in the Online Supplement), indicating rank-order stability between consecutive assessments. Cross-lagged schema effects should be interpreted as predicting residual change beyond this autoregressive component, with schema-to-autoregressive ratios ranging from 0.07 to 0.23, consistent with typical cross-lagged effect magnitudes in the literature. Interpretation of effects was based on benchmarks for cross lagged effects set by Orth and colleagues (.03 (small), .07 (medium), and .12 (large); Orth et al., 2024).

Variance inflation factors for a fully specified model indicated no problematic multicollinearity (all VIFs < 2.25). Baseline correlations among the four BCSS subscales were low to moderate (|r| = .11–.49). Residual diagnostics indicated adequate model assumptions (see Online Supplement).

### Psychosis Symptom-Schema Associations

Of 24 schema-symptom pairs tested, 12 showed significant prospective associations after FDR correction: eight unidirectional (schema predicting symptom) and four bidirectional. Schema-outcome analyses can be found in **Table 2** and are visualized in **Figure 1**; model specifics are visualized in **Figure 2**.

**Figure 1.**
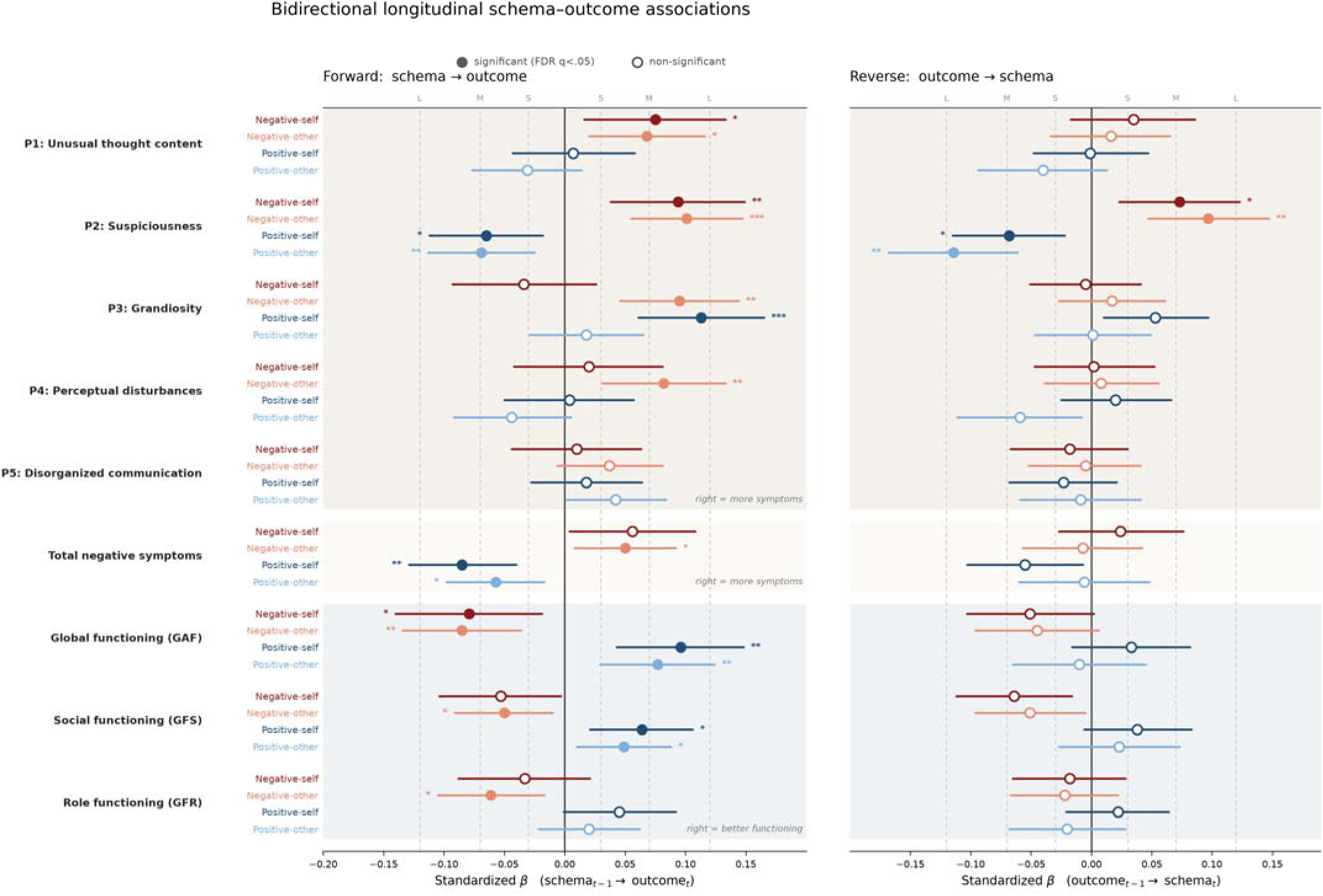
Bidirectional longitudinal schema-outcome associations. Standardized coefficients (β, 95% CI) from time-lagged mixed-effects models. **Left panel (Forward):** effect of each Brief Core Schema Scales (BCSS) subscale at the prior visit on the outcome at the following visit (schema_t−1_ → outcome_t_). **Right panel (Reverse):** effect of each outcome at the prior visit on the subscale at the following visit (outcome_t−1_ → schema_t_). Both models adjust for the autoregressive effect of the dependent variable and time varying depression. The four BCSS subscales are coded by color (negative-self, negative-other, positive-self, positive-other) outcomes are grouped into positive symptoms (SOPS P1–P5), total negative symptoms, and functioning (global [GAF], social [GFS], and role [GFR]). Filled markers are significant after Benjamini–Hochberg FDR correction (q < .05); open markers are non-significant. Asterisks denote q-value (*q<.05, **q<.01, ***q<.001). Dashed vertical lines mark small/medium/large effect-size benchmarks (Orth et al., 2024; |β| = .03/.07/.12).

**Figure 2.**
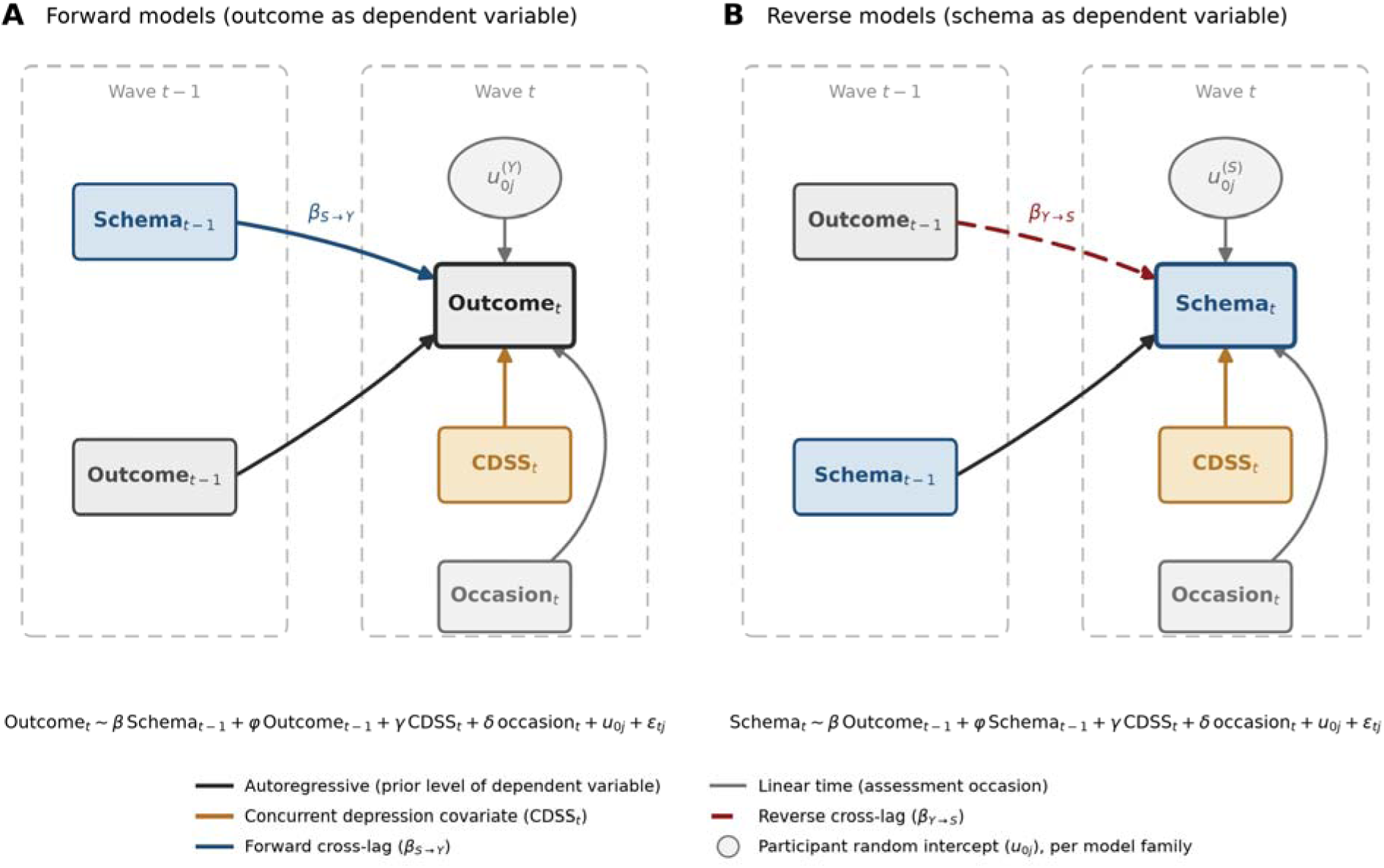
Schematic of the time-lagged mixed-effects models. Each schema–outcome pair was estimated as a forward model (A; schema at t−1 predicting outcome at t) and reverse model (B; outcome at t−1 predicting schema at t), each adjusting for the autoregressive effect of the dependent variable, concurrent depressive symptoms (CDSS), and linear time, with a participant random intercept. Cross-lagged and autoregressive coefficients pool the four t−1→t transitions.

**Table 2.**
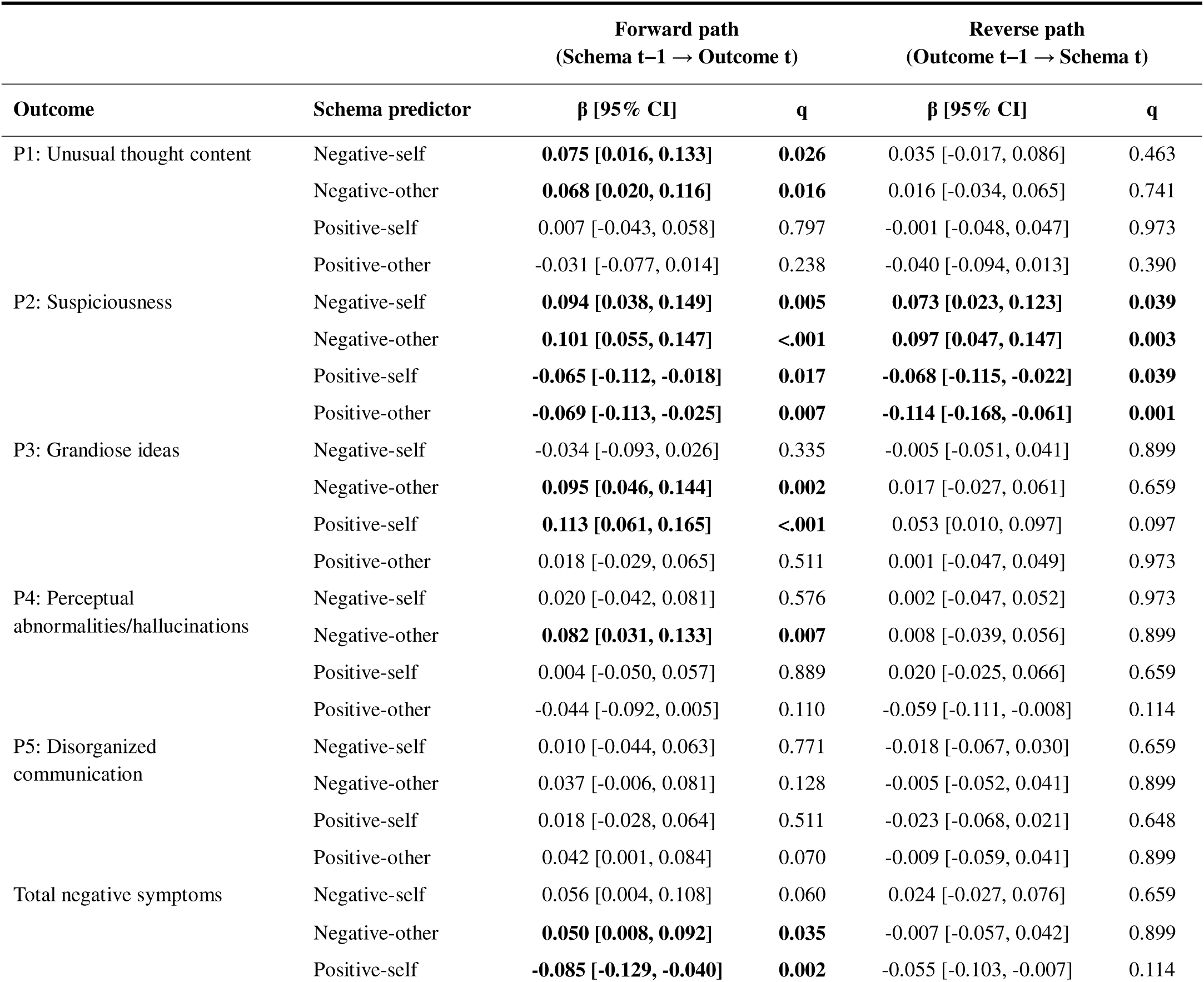
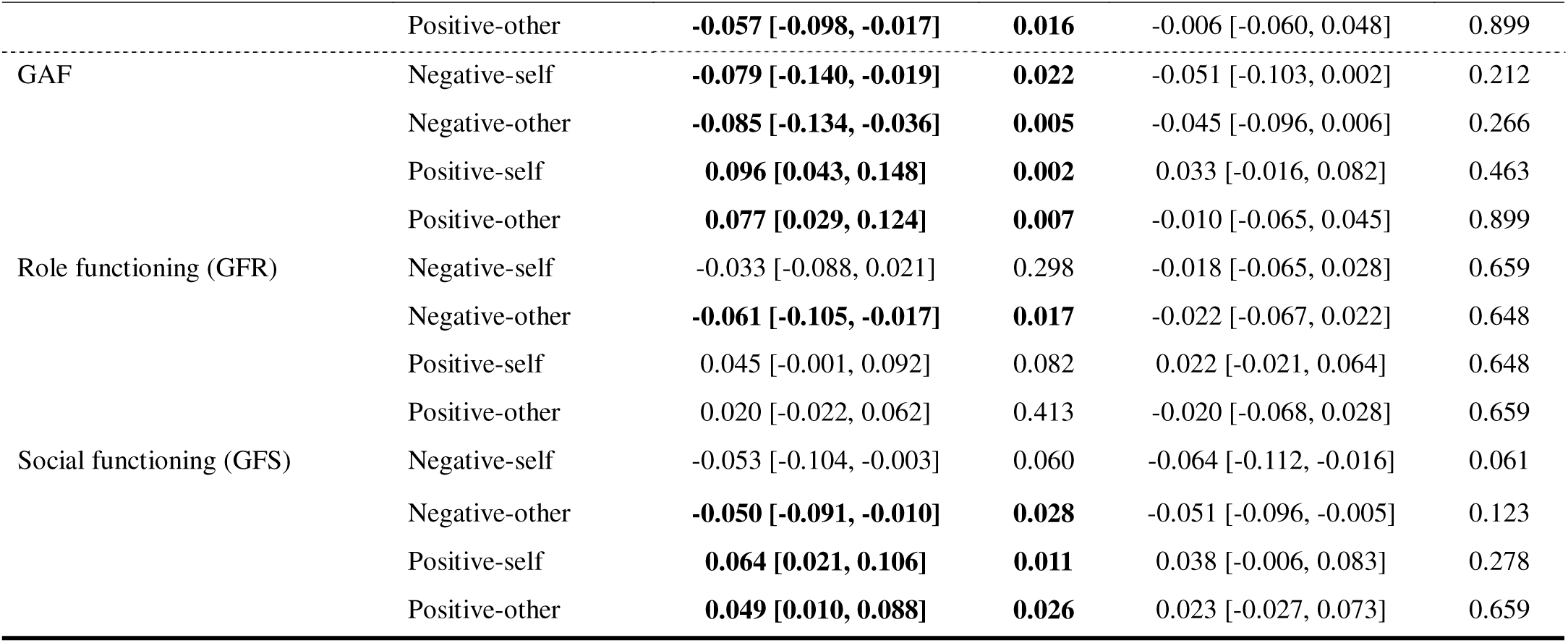
Bidirectional longitudinal schema-outcome associations.

Suspiciousness (P2) displayed the highest rate of schema-symptom relationships and bidirectional symptom-schema associations. All four schema subscales were significantly prospectively associated with suspiciousness, and all four relationships emerged as bidirectional such that suspiciousness was also prospectively associated with subsequent schema levels.

Negative schemas were prospectively associated with increased suspiciousness (negative-other: β = .101, q < .001; negative-self: β = .094, q = .005), while positive schemas appeared protective (positive-other: β = –.069, q = .007; positive-self: β = –.065, q = .017). In the reverse direction, suspiciousness at each timepoint was associated with increases in negative schemas (negative-other: β = .097, q = .003; negative-self: β = .073, q = .039) and decreases in positive schemas at the next assessment (positive-other: β = –.114, q = .001; positive-self: β = –.068, q = .039).

Grandiose ideas (P3) carried two significant forward paths. Both positive beliefs about the self (β=.113, q<0.001) and negative beliefs about others (β=0.095, q=.002) were prospectively associated with increased grandiose ideas at the subsequent timepoint. Notably, this was the only positive symptom domain wherein positive beliefs were not protective of subsequent symptom severity and actively conferred risk.

Unusual thought content/delusional ideas (P1) were prospectively associated with negative-self (β = .075, q = .026) and negative-other schemas (β = .068, q = .016), with no significant reverse paths. Perceptual abnormalities (P4) were prospectively associated with negative-other schemas (β = .082, q = .007), again with no significant reverse paths. Disorganized communication did not possess any forward, reverse, or bidirectional schema-symptom relationships.

Total negative symptoms produced three forward paths with no significant reverse effects. Both positive beliefs about oneself (β = –.085, q = .002) and positive beliefs about others (β = –.057, q = .016) were protective. Conversely, negative-other schemas were prospectively associated with greater negative symptoms at the subsequent measurement, with a smaller effect (β = .050, q = .035).

### Functioning-Schema Associations

Of 12 schema-functioning pairs tested, eight showed significant unidirectional prospective associations from schema to functioning level after FDR correction; these were the only significant associations. Global functioning (GAF) was broadly influenced by cognitive schemas, with all subscales significantly prospectively associated with subsequent GAF scores. As expected, positive schemas were prospectively associated with improved functioning over time (positive-self: β = .096, q = .002; positive-other: β = .077, q = .007), whereas negative schemas were associated with gradual decline (negative-other: β = –.085, q = .005; negative-self: β = – .079, q = .022).

Social functioning showed a similar pattern with smaller effects. Positive schemas were prospectively associated with greater subsequent social functioning (positive-self: β = .064, q = .011; positive-other: β = .049, q = .026), while negative-other schemas were associated with poorer performance (β = –.050, q = .028).

Role functioning appeared the least sensitive to schema influences, with only negative-other schemas producing a small to moderate effect (β = –.061, q = .017). All functioning effects were robust to the removal of the depression covariate.

### Sensitivity Analyses

Results were robust to multiple methodological checks (see **Online Supplement** for further details). All 20 significant paths remained significant when site was included as a crossed random effect (mean β change = 8.1%; mean site ICC = .023). Schema effects were insensitive to the inclusion of random slopes for time and to quadratic time specifications. Effects were also unchanged when visit number was replaced with participants’ observed elapsed time in months (all 20 paths remained significant; mean β change = 1.1%; Table S8). An attrition analysis indicated that longitudinal completion was unrelated to baseline symptom or schema severity supporting a missing-at-random assumption for the maximum-likelihood models **(Table S1**).

When the depression covariate was removed, all 20 forward effects remained significant (mean β change = 5.5%). However, the self-schema effects on suspiciousness were partially attenuated when depression was controlled (negative-self: 28.9% reduction; positive-self: 39.2% reduction), suggesting these prospective relationships possessed some shared variance with depressive symptoms. This is consistent with shared affective vulnerability rather than full confounding, as both paths remained significant after adjustment.

## Discussion

In a sample of CHR youth, observed up to five times across two years, both positive and negative core schemas were prospectively associated with changes in positive and negative symptoms, alongside general, social, and role functioning. Main findings were 1) negativistic cognitive schemas were prospectively associated with nearly all psychosis symptom and functioning domains, with moderate effects on attenuated positive symptom severity and functioning; 2) individual positive symptom domains possessed unique symptom and schema relationships, including bidirectional relationships with suspiciousness and unique relationships between positive schemas and grandiose ideas; 3) negative beliefs about others were prospectively associated with nearly all outcome variables; and 4) relationships between schemas and outcomes were largely independent of depressive symptoms.

These findings provide longitudinal support for cognitive models of psychosis, which position negative core schemas as vulnerabilities shaping the appraisal of neutral stimuli and emergent anomalous experiences (Garety et al., 2007, 2001; Morrison, 2001). These models posit that negative self and other schemas bias interpretation toward threatening, externally attributed explanations. Our forward paths from negative-self and negative-other schemas to unusual thought content, suspiciousness, and perceptual abnormalities are consistent with this account and extend it by demonstrating that schema fluctuations are prospectively associated with residual symptom change in a large CHR cohort (Jorovat et al., 2025; Stowkowy et al., 2016). Further, the bidirectional associations between all four schema subscales and suspiciousness empirically instantiate Freeman et al.’s (2002) threat-anticipation model wherein persecutory ideation reinforces threat-congruent beliefs about others.

Our findings prospectively support longitudinal formulations of negative schemas, affect, and psychosis symptomatology (Benrimoh et al., 2025; Georgiades et al., 2023; Gurnani and Georgiades, 2025). Self-schema effects on suspiciousness were partially attenuated by adjusting for concurrent depression, while negative-other effects were unchanged, replicating the meta-analytic finding that negative-other schemas operate independent of mood fluctuations (Jorovat et al., 2025). Further, the reverse path from suspiciousness to negative-other schemas supports the notion that real and perceived threatening social experiences, such as bullying, may shape general beliefs about others (Jorovat et al., 2025; Stowkowy and Addington, 2012). The paths between negative-other schemas and social functioning and negative symptoms extend these models to the social withdrawal often observed amongst CHR youth (Georgiades et al., 2023). Given that social stress and adversity were not modeled, future studies should consider including these variables as time-varying predictors.

The present study substantially extends prior core schema research in CHR populations. Where previous cross-sectional studies characterized schemas as stable trait-like features (Jorovat et al., 2025; Stowkowy et al., 2016), our analyses associate fluctuations in schemas with meaningful changes in psychosis symptoms and functioning over time. Similarly, we build upon previous preliminary longitudinal findings with a substantially larger sample, longer follow-up period, and models that disaggregate schemas by valence and symptoms by domain, identifying notably larger effects (Cowan et al., 2019; Patton et al., 2022). This recharacterizes schemas from static risk markers to dynamic, potentially modifiable processes with clinical significance.

Our findings also highlight novel dimensions the cognitive model underspecified. While the Garety model emphasizes vulnerability processes with little attention to resilience, our analyses reveal positive schemas as protective against suspiciousness, negative symptoms, and functional decline, effects robust to depression control. Similarly, grandiose ideas represented a notable departure from the model’s default pathway, as positive schemas’ association with future grandiose ideas suggests that grandiosity may arise through inflated self-appraisal rather than threat-based appraisal mechanisms, mirroring findings in psychotic disorder populations (Jorovat et al., 2025; Smith et al., 2006) and providing further evidence for symptom-specific elaboration within cognitive frameworks.

These findings also mirror predictive-processing accounts of psychosis (Sterzer et al., 2018). In these accounts, core schemas function as high-level priors: strongly held expectations about the self and others that shape the interpretation of ambiguous social and perceptual input (Corlett et al., 2019; Petrovic and Sterzer, 2023). Forward schema-symptom relationships suggest overweighted negative-self and negative-other priors would bias inference towards threatening interpretations, recharacterizing neutral sensory input into persecutory and delusional phenomena (Powers et al., 2017). This framework is especially supported by the bidirectional relationships observed between schemas and suspiciousness, as a threat-congruent prior yields threat-congruent percepts, which subsequently reinforce the prior, a self-confirming loop of circular inference implicated in both the formation and the consolidation of persecutory beliefs (Jardri and Denève, 2013; Reed et al., 2020). Conversely, well-calibrated positive schemas may act as stable, benign priors that constrain aberrant salience and inference. These findings suggest that computational tasks, which measure how shifts in self/other beliefs influence threat perception, may further clarify the nature of emergent suspiciousness in CHR youth.

Prospective associations with both psychosis symptom severity and functioning highlight core schemas as a potentially untapped treatment target for youth at CHR. Cognitive behavioral therapies used for CHR youth (Broekhuijse et al., 2026; French, 2004) orient towards reappraising anomalous experiences and give core schemas less attention (Akers et al., 2025). However, the eight forward paths between negative-other schemas and clinical outcomes suggest that they are a strong treatment target. While traditional cognitive behavioral therapies predominantly focus on restructuring negative schemas, positive schemas appeared protective against suspiciousness, negative symptoms, and functional decline, signaling the cultivation of positive schemas as a viable independent target (Jorovat et al., 2025). Grandiose ideas were the exception to this finding, as positive schemas preceded symptom increases; reality testing may be a superior option in this case. Our moderate effect sizes support schema change as a proximal trial outcome, particularly with regards to functioning, which existing cognitive behavioral therapies have not moved in this population (Zheng et al., 2022). Future research should identify additional psychosocial mechanisms of change relevant to CHR youth and conduct pilot trials testing the efficacy of schema-focused intervention.

Several limitations should temper interpretation of results. Primarily, time-lagged models, while stronger than cross-sectional work, do not firmly establish causation. Second, there was notable missing data on the schema measures. However, an attrition analysis indicated that participants who did and did not provide longitudinal schema data did not differ on baseline symptoms or schema severity, and baseline severity did not predict the number of schema assessments contributed (**Table S1**). Missingness was therefore consistent with a missing-at-random process, under which the maximum-likelihood estimator remains unbiased, though data missing not at random cannot be fully excluded. Third, although intraclass correlations are reported for all variables and adding random slopes for time left estimates essentially unchanged (Table S5), our lagged coefficients combine within- and between-person information rather than isolating within-person dynamics. A random-intercept cross-lagged panel model would separate these more cleanly, and replication in a cohort with additional assessment waves would make that specification viable. Fourth, the BCSS is a self-report measure of schemas, which may fail to capture the complex nature of intrapersonal beliefs. Fifth, while autoregressive controls account for symptom levels at previous timepoints, there still may be construct overlap between negative-other schemas and suspiciousness. However, negative-other items on the BCSS measure generalized beliefs about others’ character (e.g., "other people are hostile"), whereas the SIPS suspiciousness item is a clinician-rated symptom domain defined by self-referential beliefs that others intend harm to the respondent specifically. Thus, the constructs are conceptually and methodologically distinct, and their prospective association reflects the schema-to-symptom pathway rather than item redundancy.

## Conclusion

Our study provides novel evidence supporting the role of core schemas in psychosis symptom and functioning changes amongst youth at CHR. Results extend the evidence for the cognitive model of psychosis into the CHR state, a finding which should be replicated and expanded upon in future research. Symptom specific temporal associations between attenuated positive symptoms and both positive and negative schemas highlight their potential treatment value. The identification of additional mechanisms of change in CHR youth will be necessary to develop targeted and efficacious psychosocial interventions for this population.

## Data availability

Data will be made available on request. Analysis code is available at https://github.com/abroekhuijse/Core-Schemas-CHR-Analytic-Code.

## Generative AI Statement

During the preparation of this work, the author(s) used Claude Opus 5.1 in order to ensure all citations included in the text were referenced in the reference list. After using this tool/service, the author(s) reviewed and edited the content as needed and take(s) full responsibility for the content of the published article.

## CRediT author contribution

**Alexander Broekhuijse**: Conceptualization, Data curation, Formal analysis, Methodology, Project administration, Software, Validation, Visualization, Writing – original draft, Writing – review & editing. **Abhishek Saxena**: Conceptualization, Writing – review & editing. **Catalina Mourgues-Codern**: Conceptualization, Writing – review & editing. **Hesham Mukhtar**: Writing – review & editing. **Jean Addington**: Funding acquisition, Investigation, Resources, Writing – review & editing. **Carrie E. Bearden**: Funding acquisition, Investigation, Resources, Writing – review & editing. **Kristin S. Cadenhead**: Funding acquisition, Investigation, Resources, Writing – review & editing. **Daniel H. Mathalon**: Funding acquisition, Investigation, Resources, Writing – review & editing. **Thomas H. McGlashan**: Funding acquisition, Investigation, Resources, Writing – review & editing**. Diana O. Perkins**: Funding acquisition, Investigation, Resources, Writing – review & editing. **Elaine F. Walker**: Funding acquisition, Investigation, Resources, Writing – review & editing. **Tyrone D. Cannon**: Funding acquisition, Investigation, Resources, Writing – review & editing. **Scott W. Woods**: Conceptualization, Funding acquisition, Investigation, Resources, Supervision, Writing – review & editing. **Albert R. Powers III**: Conceptualization, Supervision, Writing – review & editing. **Emily A. Farina**: Conceptualization, Supervision, Writing – review & editing.

## Supporting information

Prospective Core Schemas Online Supplement

