## Supplementary material for "Prospective Bidirectional Associations Between Core Schemas, Attenuated Psychotic Symptoms, and Functioning in Youth at Clinical High Risk for Psychosis": Prospective Core Schemas Online Supplement

Analytic code for primary and supplemental analyses can be accessed at: <https://github.com/abroekhuijse/Core-Schemas-CHR-Analytic-Code>

**Sample Characteristics**

**Table S1**
Baseline Characteristics by Longitudinal Completion Status

| **Measure** | **Completers (N=275) M (SD)** | **Non-completers (N=489) M (SD)** | **t** | **df** | **p** | **d** |
| --- | --- | --- | --- | --- | --- | --- |
| Suspiciousness (P2) | 2.65 (1.49) | 2.83 (1.52) | −1.62 | 576 | .106 | −0.12 |
| Negative-other schema | 8.29 (6.09) | 7.60 (6.11) | 1.46 | 574 | .146 | 0.11 |
| Negative-self schema | 6.60 (5.52) | 6.38 (5.66) | 0.51 | 584 | .613 | 0.04 |
| Global functioning (GAF) | 49.82 (11.59) | 47.57 (10.17) | 2.68 | 511 | .008 | 0.21 |
| Depression (CDSS) | 5.91 (4.90) | 5.76 (4.67) | 0.41 | 558 | .683 | 0.03 |

*Note. Completers contributed BCSS ratings at ≥ 4 of 5 visits (n = 275); non-completers at ≤ 3 visits (n = 489). Baseline groups were compared with Welch t tests; d = Cohen's d. Groups did not differ on baseline symptoms, schemas, or depression; completers had slightly higher baseline global functioning (GAF; d = 0.21). In a regression predicting the number of BCSS waves from all five baseline measures, only GAF was a significant predictor (β = .147, p = .014; model R² = .017). Retention was thus unrelated to baseline symptom or schema severity, supporting the missing-at-random assumption underlying the mixed-effects models.*

**Model Diagnostics**

**Figure S1**
*Residual Diagnostics for a Representative Model (Suspiciousness [P2] Regressed on Prior Negative-Other Schema)*

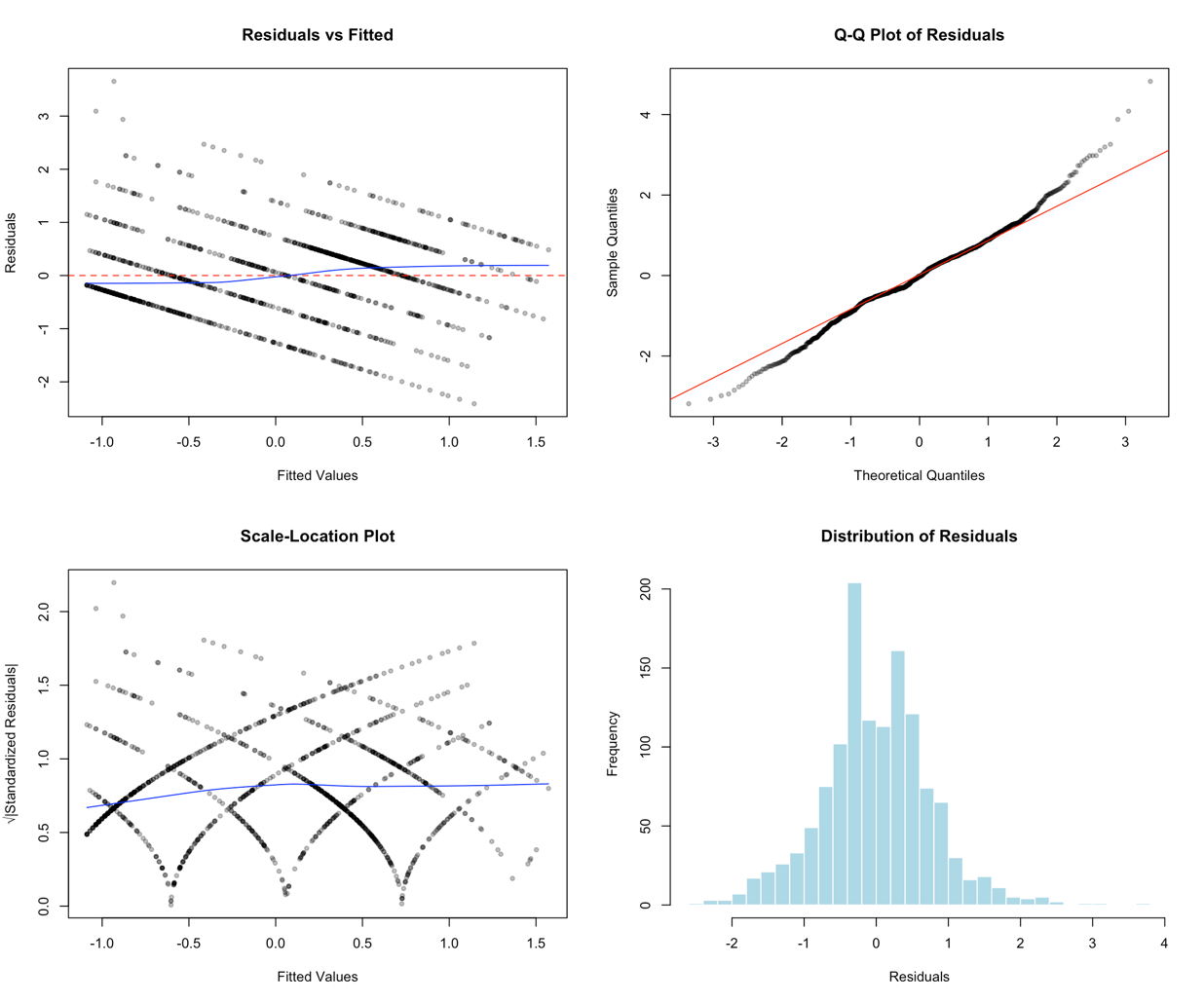

*Note.* Panels show residuals vs. fitted values, a normal Q-Q plot, a scale-location plot, and the residual distribution. Shapiro-Wilk for this model: *W* = .989, *p* = 1.7 × 10⁻⁸. With N ≈ 1,280, the test flags trivial departures from normality; the Q-Q plot indicates approximate normality. Banding in the residual plots reflects the discrete (integer) SIPS rating scale.

**Table S2**
*Residual Diagnostics Across All Significant Forward-Path (Schema → Outcome) Models*

| **Outcome** | **Schema** | **SD** | **Skew** | **Kurtosis** | **Shapiro W** | **Shapiro p** |
| --- | --- | --- | --- | --- | --- | --- |
| P1 | Negative-self | 0.653 | −0.128 | 3.316 | 0.995 | < .001 |
| P1 | Negative-other | 0.661 | −0.102 | 3.300 | 0.995 | < .001 |
| P2 | Negative-self | 0.758 | 0.162 | 4.049 | 0.988 | < .001 |
| P2 | Negative-other | 0.756 | 0.168 | 4.090 | 0.989 | < .001 |
| P2 | Positive-self | 0.758 | 0.154 | 4.057 | 0.988 | < .001 |
| P2 | Positive-other | 0.758 | 0.173 | 4.089 | 0.988 | < .001 |
| P3 | Negative-other | 0.687 | 0.831 | 5.066 | 0.908 | < .001 |
| P3 | Positive-self | 0.675 | 0.850 | 5.106 | 0.911 | < .001 |
| P4 | Negative-other | 0.696 | −0.093 | 2.920 | 0.994 | < .001 |
| Neg. symptoms | Negative-other | 0.706 | 0.195 | 4.138 | 0.988 | < .001 |
| Neg. symptoms | Positive-self | 0.701 | 0.174 | 4.135 | 0.987 | < .001 |
| Neg. symptoms | Positive-other | 0.703 | 0.183 | 4.149 | 0.988 | < .001 |
| GAF | Negative-self | 0.666 | −0.009 | 4.890 | 0.978 | < .001 |
| GAF | Negative-other | 0.684 | −0.034 | 4.923 | 0.978 | < .001 |
| GAF | Positive-self | 0.668 | −0.001 | 4.811 | 0.978 | < .001 |
| GAF | Positive-other | 0.665 | −0.026 | 4.854 | 0.978 | < .001 |
| GF:R | Negative-other | 0.748 | −0.550 | 4.697 | 0.964 | < .001 |
| GF:S | Negative-other | 0.685 | −0.176 | 4.414 | 0.987 | < .001 |
| GF:S | Positive-self | 0.682 | −0.140 | 4.423 | 0.986 | < .001 |
| GF:S | Positive-other | 0.684 | −0.145 | 4.390 | 0.987 | < .001 |

*Note.* Residual means were 0 by construction and are omitted. Shapiro-Wilk *p* < .001 for all models, expected at this sample size; SD, skewness, and kurtosis indicate residuals approximately centered and near-mesokurtic, supporting adequate model assumptions.

**Multicolinearity**

**Table S3**
*Variance Inflation Factors for the Fully Specified Suspiciousness (P2) Model*

| **Predictor** | **VIF** |
| --- | --- |
| Prior suspiciousness (P2) | 1.40 |
| Prior negative-self schema | 2.24 |
| Prior negative-other schema | 1.49 |
| Prior positive-self schema | 1.80 |
| Prior positive-other schema | 1.47 |
| Prior depression (CDSS) | 1.80 |
| Assessment wave | 1.10 |

*Note.* Model: P2 ~ all four schemas + prior P2 + prior depression + assessment wave. All VIFs < 2.25; values > 5 indicate problematic multicollinearity.

**Table S4**
*Baseline Intercorrelations Among BCSS Subscales*

|  | **Negative-self** | **Negative-other** | **Positive-self** | **Positive-other** |
| --- | --- | --- | --- | --- |
| **Negative-self** | — | .478 | −.482 | −.214 |
| **Negative-other** | .478 | — | −.110 | −.122 |
| **Positive-self** | −.482 | −.110 | — | .491 |
| **Positive-other** | −.214 | −.122 | .491 | — |

*Note.* Pearson correlations at baseline. Inter-subscale associations were low to moderate (|r| = .11–.49), consistent with distinct schema dimensions.

**Sensitivity Analyses**

**Table S5**
*Random-Intercept vs. Random-Slope Specifications*

| **Outcome** | **Schema** | **AIC (RI)** | **AIC (RS)** | **β (RI)** | **β (RS)** | **ΔAIC** | **Δβ** |
| --- | --- | --- | --- | --- | --- | --- | --- |
| P1 | Negative-self | 2962.18 | 2952.89 | .075 | .072 | −9.28 | .002 |
| P1 | Negative-other | 2963.37 | 2955.83 | .068 | .062 | −7.53 | .006 |
| P2 | Negative-self | 2953.52 | 2954.26 | .094 | .091 | 0.74 | .003 |
| P2 | Negative-other | 2942.86 | 2943.72 | .101 | .098 | 0.86 | .003 |
| P2 | Positive-self | 2961.43 | 2960.31 | −.065 | −.069 | −1.12 | .004 |
| P2 | Positive-other | 2957.33 | 2957.36 | −.069 | −.068 | 0.03 | .001 |
| P3 | Negative-other | 3026.23 | 3026.68 | .095 | .092 | 0.45 | .003 |
| P3 | Positive-self | 3030.06 | 3030.27 | .113 | .110 | 0.22 | .003 |
| P4 | Negative-other | 3104.40 | 3103.44 | .082 | .082 | −0.96 | .001 |
| Neg. symptoms | Negative-other | 2780.31 | 2776.40 | .050 | .050 | −3.91 | .001 |
| Neg. symptoms | Positive-self | 2773.52 | 2768.09 | −.084 | −.091 | −5.42 | .007 |
| Neg. symptoms | Positive-other | 2786.45 | 2782.73 | −.057 | −.057 | −3.71 | .000 |
| GAF | Negative-self | 3035.85 | 3037.14 | −.079 | −.077 | 1.29 | .003 |
| GAF | Negative-other | 3027.04 | 3027.79 | −.085 | −.085 | 0.75 | .000 |
| GAF | Positive-self | 3041.87 | 3041.46 | .095 | .098 | −0.41 | .002 |
| GAF | Positive-other | 3040.72 | 3042.11 | .076 | .073 | 1.39 | .003 |
| GF:R | Negative-other | 2900.34 | 2904.31 | −.061 | −.061 | 3.97 | .000 |
| GF:S | Negative-other | 2678.99 | 2682.99 | −.050 | −.050 | 4.00 | .000 |
| GF:S | Positive-self | 2677.27 | 2681.22 | .064 | .064 | 3.94 | .000 |
| GF:S | Positive-other | 2683.58 | 2687.57 | .049 | .049 | 3.98 | .000 |

*Note. Adding a random slope for time changed schema coefficients negligibly (|Δβ| ≤ .007) and did not consistently improve fit (the random-slope model lowered AIC in 8 of 20 models). Illustratively, for P2 ~ prior negative-other, the likelihood-ratio test was non-significant (χ² = 3.14, df = 2, p = .208). ΔAIC < 0 favors the random-slope model.*

**Table S6**
*Linear vs. Quadratic Specification of Time*

| **Model** | **AIC (linear)** | **AIC (quadratic)** | **LRT χ²** | **p** | **β (linear)** | **β (quadratic)** |
| --- | --- | --- | --- | --- | --- | --- |
| P1 ~ prior negative-self | 2962.18 | 2963.14 | 1.04 | .309 | .075 | .074 |
| P1 ~ prior negative-other | 2963.37 | 2964.24 | 1.12 | .289 | .068 | .067 |
| P2 ~ prior negative-self | 2953.52 | 2953.79 | 1.73 | .189 | .094 | .093 |
| P2 ~ prior negative-other | 2942.86 | 2942.98 | 1.88 | .170 | .101 | .100 |
| P2 ~ prior positive-self | 2961.43 | 2961.49 | 1.94 | .163 | −.065 | −.064 |
| P2 ~ prior positive-other | 2957.33 | 2957.32 | 2.01 | .156 | −.069 | −.068 |
| P3 ~ prior negative-other | 3026.23 | 3028.17 | 0.07 | .798 | .095 | .095 |
| P3 ~ prior positive-self | 3030.06 | 3032.01 | 0.04 | .839 | .113 | .113 |
| P4 ~ prior negative-other | 3104.40 | 3105.62 | 0.78 | .377 | .082 | .081 |
| Neg. symptoms ~ prior negative-other | 2780.31 | 2781.79 | 0.52 | .473 | .050 | .050 |
| Neg. symptoms ~ prior positive-self | 2773.52 | 2775.10 | 0.42 | .517 | −.084 | −.084 |
| Neg. symptoms ~ prior positive-other | 2786.45 | 2788.02 | 0.42 | .516 | −.057 | −.057 |
| GAF ~ prior negative-self | 3035.85 | 3034.55 | 3.30 | .069 | −.079 | −.078 |
| GAF ~ prior negative-other | 3027.04 | 3025.57 | 3.47 | .062 | −.085 | −.084 |
| GAF ~ prior positive-self | 3041.87 | 3040.76 | 3.11 | .078 | .095 | .094 |
| GAF ~ prior positive-other | 3040.72 | 3039.84 | 2.87 | .090 | .076 | .074 |
| GF:R ~ prior negative-other | 2900.34 | 2901.89 | 0.45 | .502 | −.061 | −.061 |
| GF:S ~ prior negative-other | 2678.99 | 2680.81 | 0.18 | .671 | −.050 | −.050 |
| GF:S ~ prior positive-self | 2677.27 | 2679.10 | 0.18 | .674 | .064 | .064 |
| GF:S ~ prior positive-other | 2683.58 | 2685.40 | 0.18 | .668 | .049 | .049 |

*Note. A quadratic time term did not improve fit for any of the 20 significant forward-path models (all likelihood-ratio p > .05) and left schema effects essentially unchanged (|Δβ| ≤ .002). Models compared by likelihood-ratio test under maximum likelihood.*

**Table S7**
*Schema Effects With vs. Without the Depression Covariate (All 20 Significant Forward Paths)*

| **Outcome** | **Schema** | **β (with)** | **p (with)** | **β (no dep.)** | **p (no dep.)** | **Δ %** | **Interpretation** |
| --- | --- | --- | --- | --- | --- | --- | --- |
| P1 | Negative-self | .075 | .012 | .074 | .002 | −0.5 | Robust |
| P1 | Negative-other | .068 | .006 | .074 | .002 | 8.0 | Robust |
| P2 | Negative-self | .094 | .001 | .121 | < .001 | 28.9 | **Partial mediation** |
| P2 | Negative-other | .101 | < .001 | .116 | < .001 | 15.2 | Robust |
| P2 | Positive-self | −.065 | .007 | −.090 | < .001 | 39.2 | **Partial mediation** |
| P2 | Positive-other | −.069 | .002 | −.079 | < .001 | 13.7 | Robust |
| P3 | Negative-other | .095 | < .001 | .082 | .001 | −13.6 | Robust |
| P3 | Positive-self | .113 | < .001 | .098 | < .001 | −13.6 | Robust |
| P4 | Negative-other | .082 | .002 | .082 | .001 | 0.1 | Robust |
| Neg. symptoms | Negative-other | .050 | .020 | .050 | .014 | −0.1 | Robust |
| Neg. symptoms | Positive-self | −.085 | < .001 | −.076 | < .001 | −9.9 | Robust |
| Neg. symptoms | Positive-other | −.057 | .006 | −.054 | .008 | −5.4 | Robust |
| GAF | Negative-self | −.079 | .010 | −.082 | .002 | 2.7 | Robust |
| GAF | Negative-other | −.085 | .001 | −.090 | < .001 | 5.5 | Robust |
| GAF | Positive-self | .096 | < .001 | .099 | < .001 | 3.1 | Robust |
| GAF | Positive-other | .077 | .002 | .079 | .001 | 3.1 | Robust |
| GF:R | Negative-other | −.061 | .006 | −.062 | .003 | 2.4 | Robust |
| GF:S | Negative-other | −.050 | .015 | −.059 | .003 | 18.0 | Robust |
| GF:S | Positive-self | .064 | .003 | .069 | .001 | 8.4 | Robust |
| GF:S | Positive-other | .049 | .013 | .051 | .009 | 4.5 | Robust |

*Note.* All 20 effects remained significant without the depression covariate (mean |Δβ| = 5.5%). The self-schema → suspiciousness paths attenuated most when depression was controlled (negative-self 28.9%; positive-self 39.2%), consistent with partially shared affective variance rather than confounding. Δ % = percent change in β when depression is removed.

**Table S8**
Robustness to Observed Elapsed Time (Months) as the Time Metric

| **Outcome** | **Schema** | **β (visit no.)** | **β (months)** | **p (months)** | **Δ %** |
| --- | --- | --- | --- | --- | --- |
| P1 | Negative-self | .075 | .075 | .011 | 0.9 |
| P1 | Negative-other | .068 | .070 | .005 | 2.1 |
| P2 | Negative-self | .094 | .093 | .001 | −0.5 |
| P2 | Negative-other | .101 | .103 | < .001 | 1.6 |
| P2 | Positive-self | −.065 | −.065 | .006 | 0.8 |
| P2 | Positive-other | −.069 | −.070 | .002 | 1.7 |
| P3 | Negative-other | .095 | .096 | < .001 | 0.8 |
| P3 | Positive-self | .113 | .113 | < .001 | 0.0 |
| P4 | Negative-other | .082 | .082 | .002 | 0.6 |
| Neg. symptoms | Negative-other | .050 | .052 | .017 | 2.7 |
| Neg. symptoms | Positive-self | −.085 | −.085 | < .001 | 0.7 |
| Neg. symptoms | Positive-other | −.057 | −.058 | .005 | 1.6 |
| GAF | Negative-self | −.079 | −.079 | .010 | −0.1 |
| GAF | Negative-other | −.085 | −.086 | < .001 | 1.4 |
| GAF | Positive-self | .096 | .096 | < .001 | 0.5 |
| GAF | Positive-other | .077 | .078 | .001 | 1.2 |
| GF:R | Negative-other | −.061 | −.063 | .005 | 2.5 |
| GF:S | Negative-other | −.050 | −.051 | .013 | 1.9 |
| GF:S | Positive-self | .064 | .064 | .003 | 0.2 |
| GF:S | Positive-other | .049 | .049 | .012 | 0.8 |

*Note. Each significant forward-path model was refit replacing visit number with the actual elapsed time since baseline (in months; median assessment spacing = 0, 5.9, 12.1, 17.7, and 24.0 months), standardized. All 20 paths remained significant with the observed-time metric, and schema coefficients were essentially unchanged from the primary models (mean |Δβ| = 1.1%, maximum 2.7%). β (visit no.) reproduces the primary model; Δ % = percent change in the schema coefficient when observed elapsed time replaces visit number. This indicates the linear-time specification is not an artifact of treating visits as equally spaced.*

**Table S9**
*Site as a Crossed Random Effect*

| **Outcome** | **Schema** | **β (no site)** | **β (+ site)** | **p (+ site)** | **Δ %** | **Site ICC** |
| --- | --- | --- | --- | --- | --- | --- |
| P1 | Negative-self | .075 | .075 | .012 | 0.5 | .002 |
| P1 | Negative-other | .068 | .068 | .006 | −0.2 | .002 |
| P2 | Negative-self | .094 | .114 | < .001 | 21.1 | .031 |
| P2 | Negative-other | .101 | .106 | < .001 | 4.7 | .026 |
| P2 | Positive-self | −.065 | −.076 | .002 | 16.6 | .027 |
| P2 | Positive-other | −.069 | −.072 | .001 | 4.3 | .025 |
| P3 | Negative-other | .095 | .098 | < .001 | 3.4 | .008 |
| P3 | Positive-self | .113 | .111 | < .001 | −1.8 | .003 |
| P4 | Negative-other | .082 | .085 | .001 | 3.4 | .009 |
| Neg. symptoms | Negative-other | .050 | .047 | .029 | −6.5 | .013 |
| Neg. symptoms | Positive-self | −.085 | −.094 | < .001 | 11.4 | .017 |
| Neg. symptoms | Positive-other | −.057 | −.057 | .006 | 0.4 | .014 |
| GAF | Negative-self | −.079 | −.101 | < .001 | 26.7 | .053 |
| GAF | Negative-other | −.085 | −.088 | < .001 | 3.2 | .047 |
| GAF | Positive-self | .096 | .120 | < .001 | 25.0 | .055 |
| GAF | Positive-other | .077 | .091 | < .001 | 18.2 | .053 |
| GF:R | Negative-other | −.061 | −.064 | .004 | 4.7 | .011 |
| GF:S | Negative-other | −.050 | −.048 | .019 | −4.0 | .021 |
| GF:S | Positive-self | .064 | .078 | < .001 | 21.6 | .025 |
| GF:S | Positive-other | .049 | .054 | .006 | 9.8 | .023 |

*Note. Each significant forward-path model was refit with study site added as a crossed random intercept, (1 | ID) + (1 | Site), across the eight NAPLS-2 sites. All 20 paths remained significant with site included (mean β change = 8.1%; mean site ICC = .023, range .002–.055). β (no site) reproduces the primary model; Δ % = percent change in the schema coefficient when site is added. The small site ICCs indicate little between-site variance after accounting for the modeled covariates.*
